# Development and Multi-Site External Validation of a Generalizable Risk Prediction Model for Bipolar Disorder

**DOI:** 10.1101/2023.02.21.23286251

**Authors:** Colin G. Walsh, Michael A. Ripperger, Yirui Hu, Yi-han Sheu, Drew Wilimitis, Amanda B. Zheutlin, Daniel Rocha, Karmel W. Choi, Victor M. Castro, H. Lester Kirchner, Christopher F. Chabris, Lea K. Davis, Jordan W. Smoller

**Author notes:** Corresponding Author: Colin G. Walsh, 2525 West End Ave Suite 1475, Nashville, TN 37203.

## Abstract

Bipolar disorder is a leading contributor to disability, premature mortality, and suicide. Early identification of risk for bipolar disorder using generalizable predictive models trained on diverse cohorts around the United States could improve targeted assessment of high risk individuals, reduce misdiagnosis, and improve the allocation of limited mental health resources.

This observational case-control study intended to develop and validate generalizable predictive models of bipolar disorder as part of the multisite, multinational PsycheMERGE Consortium across diverse and large biobanks with linked electronic health records (EHRs) from three academic medical centers: in the Northeast (Massachusetts General Brigham), the Mid-Atlantic (Geisinger) and the Mid-South (Vanderbilt University Medical Center).

Predictive models were developed and validated with multiple algorithms at each study site: random forests, gradient boosting machines, penalized regression, including stacked ensemble learning algorithms combining them. Predictors were limited to widely available EHR-based features agnostic to a common data model including demographics, diagnostic codes, and medications. The main study outcome was bipolar disorder diagnosis as defined by the International Cohort Collection for Bipolar Disorder, 2015.

In total, the study included records for 3,529,569 patients including 12,533 cases (0.3%) of bipolar disorder. After internal and external validation, algorithms demonstrated optimal performance in their respective development sites. The stacked ensemble achieved the best combination of overall discrimination (AUC = 0.82 - 0.87) and calibration performance with positive predictive values above 5% in the highest risk quantiles at all three study sites.

In conclusion, generalizable predictive models of risk for bipolar disorder can be feasibly developed across diverse sites to enable precision medicine. Comparison of a range of machine learning methods indicated that an ensemble approach provides the best performance overall but required local retraining. These models will be disseminated via the PsycheMERGE Consortium website.

## Introduction

Bipolar disorder (BD), characterized by episodes of hypomania/mania and depression^1^, is a leading cause of disability.^2^ Rates of suicide among patients with BD are 20- to 30-fold higher than in the general population,^3^ and BD is associated with substantial premature mortality from multiple causes.^4^ The diagnosis of BD can be challenging and may require a prolonged diagnostic odyssey, averaging 6-10 years.^5–7^ Affected patients frequently present initially with a major depressive episode and are misdiagnosed with unipolar depression. Misdiagnosis may lead to inappropriate prescribing of antidepressants without mood stabilization and increased risk of switching into a manic state.^8^ Duration of untreated bipolar disorder is associated with more severe and recurrent mood episodes and more frequent suicide attempts.^9,10^

Identifying those at risk for BD might enable targeted assessment, early intervention, and more appropriate management. A recent systematic review of clinical trials to prevent bipolar disorder showed reliance on family history for risk identification.^11^ However, given BD’s multifactorial nature, most affected would not have a positive family history.^12^ In addition and unlike schizophrenia, no established prodrome exists for bipolar disorder. Newer methods for risk identification not reliant on existing clinical signs or symptoms might be of substantial value.

Longitudinal electronic health records (EHRs) coupled with predictive analytics might enable novel risk identification opportunities in BD. We have previously demonstrated that such data can produce valid diagnostic phenotyping of bipolar cases.^13,14^ Here, we extend this work to the domain of prognostication by leveraging the PsycheMERGE consortium, a national research network of EHR-linked biobanks. Using longitudinal EHRs from three major healthcare systems, we trained and validated quantitative bipolar disorder risk prediction models based on high-dimensional structured EHR data and evaluated their performance individually and when ensembled.

## Methods

### Study Settings

Participating study sites included three major academic medical centers in the United States: the Northeast (Mass General Brigham [MGB]) the Mid-South (Vanderbilt University Medical Center [VUMC]), the Mid-Atlantic (Geisinger [GHS]). Each site participates in the PsycheMERGE Consortium and has an extensive EHR repository linked to a biobank. On average, these sites each serve 1.4M patients per year and have EHR repositories of ∼2.7M patients linked via EHRs.

The methods were performed in accordance with relevant guidelines and regulations and approved by Institutional Review Boards at each participating study site: Vanderbilt University Medical Center, Geisinger Health System, Mass General Brigham.

### Outcome Definition

Cases of BD were defined by the published “Bipolar Coded-Broad” definition per Castro et al., 2015.^14^ This rule-based algorithm demonstrated high positive predictive value (0.80) using a gold-standard of semi-structured diagnostic interviews (SCID-IV) by experienced doctoral-level clinicians blind to algorithmic results. To meet “Bipolar Coded-Broad,” cases must have at least two BD diagnostic codes versions nine or ten of the International Classification of Diseases (ICD) schema with a minimum of four weeks between each code and at least two documented medications used to treat BD (e.g., lithium or valproic acid) within one year of the index BD diagnosis. To rule out patients with related disorders, we required the number of diagnostic codes for major depressive disorder (MDD), schizophrenia (SCZ), schizoaffective disorder, or organic affective syndrome (OAS) to total fewer than half the number of BD codes. Only adult patients aged 18 years and older at the time of their index BD diagnosis were included in this analysis.

All adult patients were included if they had a minimum of three documented healthcare encounters over a minimum of six months, regardless of case status.

### Predictive Modeling Approach

We compared three predictive modeling approaches that together span a range of model architectures and strategies for handling feature relationships (see “Algorithmic Details”, below). Because of prior algorithmic experience at each study site, each team validated internally one of the following with multisite external validation at the other two sites: L2-penalized regression (abbreviated here as “Ridge”) at MGB, random forests (RF) at VUMC, and gradient boosting machines (GBM) at GHS.

Internal validation was conducted at each site with a randomly selected hold-out test and the best internally performing algorithms were shared for external validation. This reciprocal validation strategy tested generalizability of each algorithm without the need for each site to train three separate algorithms in parallel.

### Feature Engineering

Variables for prediction included demographics: age (continuous), coded sex (categorical: Male, Female, and Unknown), coded race (categorical: White, Black, Asian, Other and Unknown); inpatient-administered and outpatient-prescribed medications (log-transformed counts); and diagnostic codes (log-transformed counts). Dimensionality reduction included grouping medications by their RxNorm ingredients^15^ and diagnostic codes mapped from ICD-9-CM and ICD-10-CM to Clinical Classification Software (CCS) Level 2 codes.^16^ The final feature list numbered up to ∼2500. Missing data for count variables were imputed as zeroes and categorized as unknown for race and sex (see Table 1).

**Table 1:** Baseline study participant characteristics

| Site | VUMC (%) | GHS (%) | MGB (%) |
| --- | --- | --- | --- |
| Total Patients in Study, N | 932,784 | 934,749 | 1,662,036 |
| Total Patients, Bipolar Coded-Broad, N (%) | 3,357 (0.36%) | 3,101 (0.33%) | 6,075(0.36%) |
| Internal Validation Training Set, N (%) | 746,226 (80.0%) | 701,147 (75.0%) | 997,687 (60.0%) |
| Internal Validation Testing Set, N (%) | 186,558 (20.0%) | 232,942 (25.0%) | 664,349 (40.0%) |
| Sex, Women | 535,273 (57.4%) | 524,479 (56%) | 980,586 (59%) |
| Sex, Men | 392,477 (42.6%) | 410,270 (44%) | 681,372 (41%) |
| Sex, Unknown | 34 (0.0036%) | 0 (0%) | 78 (0.0047%) |
| Coded Race, White | 701,525 (75.2%) | 881,433 (94.3%) | 1,282,679 (77%) |
| Coded Race, Black | 100,462(10.8%) | 33,025 (3.6%) | 106,362 (6.4%) |
| Coded Race, Asian | 16,055 (1.72%) | 9,129 (1.0%) | 69,651 (4.1%) |
| Coded Race, Other | 32,063 (3.43%) | 5,786 (0.6%) | 203,315 (12%) |
| Coded Race, Unknown | 82,679 (8.86%) | 5,490 (0.6%) | 29 (0%) |
| Age, Mean (Range) | 49.5 (18-89) | 52.2 (18-89) | 52.82 (18-89) |
| EHR Length in Years, median (Q1-Q3) | 7 (3-13) | 8 (3-15) | 9 (4-15) |

Records meeting “Bipolar Coded Broad” were right-censored until the day before index diagnosis to represent a useful prediction target and to prevent leakage of bipolar-related data from driving model predictions. For those not meeting bipolar disorder criteria, right-censoring occurred at the last date of a visit or first occurrence of an ICD code for BD in the EHR.

### Algorithmic Details

#### Ridge Regression

Ridge Regression^17^ is a regularized regression model that imposes shrinkage of regression coefficients to reduce multi-collinearity and model variance, and thereby increasing prediction performance. Despite the shrinkage, the regression coefficients are never shrunk to zero, and therefore all features remain in the final model. We used the widely adopted *glmnet*^18-20^ package in R for model training, using the main (first order) effects of all available features. The model was developed with 10-fold cross-validation using 60% of all data to find the best Lambda value (i.e., strength of regularization) and estimate model parameters, while the remaining 40% data were used as a hold-out test set. The 60-40 split was chosen due to a larger sample size at MGB, and the 60% training/validation split approached the limits in input data size for the *glmnet* package. Preliminary analyses showed minimal performance differences by varying the train/test ratio.

#### RF

VUMC implemented the decision-tree based RF. A commonly employed nonparametric algorithm, RF permits nonlinear relationships between predictors, samples with replacement for feature inclusion and model training, and it tolerates collinearity likely to be present in EHR data. After preliminary analyses varying the following, parameters of 200 trees, minimum node size of five, and purity for importance were used. Both RF and GBM below were trained with an 80-20% train-test split. RF were implemented using the ranger package in R.^21^

#### GBM

Boosting is an ensemble technique based on using multiple “weak learner” algorithms to train a strong one through sequential training to iteratively improve prediction accuracy. GBM is a high-performance gradient boosting framework based on decision trees capable of handling imbalanced datasets, as the boosting can strengthen the impact of the positive class (here, cases of BD). Tuning parameters included the ratio of features used, the ratio of training instances, maximum depth of trees and the learning rate. In preliminary analyses, dimensionality impacted model performance, so we selected the most prevalent medications across the GHS EHR by including those accounting for 95% of cumulative medication counts by Pareto analysis. Here, GBM were implemented in Python (package ‘lightgbm’).^22^

#### Ensembling

Ensembling is designed to improve prediction accuracy by aggregating the strengths of diverse machine learning models into a single predictive model. Here, we ensembled the three algorithms via stacking of all three algorithms at each site and evaluated performance. We combined Ridge, GBM, and RFs with a stacked ensemble trained with ten-fold cross validation and logistic regression using the three individual predictions as multivariate predictors on each site’s training set to avoid leakage of test data for the internally valid model into the ensemble.

#### Model Evaluation

Evaluation metrics included Area Under the Receiver Operating Characteristic (AUROC) and Precision-Recall Curves and Area Under the P-R Curve (AUPR). Metrics at specific risk thresholds included sensitivity/recall, specificity, positive predictive value (PPV), and number needed to screen (NNS,^23^ the reciprocal of PPV for a predictive model) Calibration was measured with Brier score and calibration slope/intercept.^24^

## Results

Study site data are shown (Table 1).

### Individual Model Performance by Site

Discrimination performance is shown for each algorithm by training site with internal validation (testing within site) denoted visually for ready comparison to external validation (testing across sites) (Table 2).

**Table 2:** Model Performance by Site. Internal Validation italicized and denoted by color. AUROC = Area Under the Receiver Operating Characteristic; AUPR = Area Under the Precision-Recall Curve

|  | AUROC | AUPR | Brier Score | Calibration Slope | Calibration Intercept |
| --- | --- | --- | --- | --- | --- |
| **VUMC** |  |  |  |  |  |
| RIDGE | 0.796 | 0.020 | 0.004 | 2.108 | 6.085 |
| RF | 0.836 | 0.046 | 0.004 | 0.946 | -0.303 |
| GBM | 0.808 | 0.025 | 0.004 | 1.011 | 0.092 |
| ENSEMBLE | 0.837 | 0.049 | 0.004 | 0.996 | -0.021 |
| **GHS** |  |  |  |  |  |
| RIDGE | 0.775 | 0.015 | 0.003 | 1.987 | 5.310 |
| RF | 0.775 | 0.015 | 0.003 | 0.625 | -2.108 |
| GBM | 0.873 | 0.054 | 0.003 | 1.085 | 0.387 |
| ENSEMBLE | 0.825 | 0.032 | 0.003 | 1.027 | 0.224 |
| **MGB** |  |  |  |  |  |
| RIDGE | 0.865 | 0.026 | 0.004 | 1.502 | 2.674 |
| RF | 0.802 | 0.019 | 0.004 | 0.726 | -1.626 |
| GBM | 0.852 | 0.033 | 0.004 | 1.000 | -0.002 |
| ENSEMBLE | 0.822 | 0.026 | 0.004 | 0.981 | -0.111 |

As shown in Table 2, models performed comparably within and across sites with a tendency for better discrimination at internal validation sites for locally trained algorithms and better calibration for GBM and Ensembles of GBM, RF and Ridge.

### Optimal thresholds and performance metrics by algorithm and site

Varying risk percentile thresholds by algorithm and by site showed specificity was closely linked to the thresholds themselves, while sensitivity (recall) and PPV tended to decrease and increase, respectively, as thresholds increased (Table 3). NPV for all algorithms above these thresholds (90% and above) was over 99%, largely because of the rarity of the outcomes in question.

**Table 3:** Model performance by risk percentile threshold

|  | Risk Percentile Cutoff | Specificity (%) | Sensitivity (%) | PPV (%) | NPV (%) |
| --- | --- | --- | --- | --- | --- |
| **Random Forest** |  |  |  |  |  |
| VUMC | *90* | *90.2* | *58.6* | *2.1* | *99.8* |
|  | *95* | *95.2* | *45.5* | *3.3* | *99.8* |
|  | *99* | *99.1* | *19.2* | *6.9* | *99.7* |
| MGB | 90 | 90.2 | 51.7 | 1.9 | 99.8 |
|  | 95 | 95.1 | 34.9 | 2.5 | 99.7 |
|  | 99 | 99 | 9.1 | 3.3 | 99.7 |
| GHS | 90 | 90.1 | 44.8 | 1.5 | 99.8 |
|  | 95 | 95.1 | 30.1 | 2 | 99.8 |
|  | 99 | 99 | 7.6 | 2.5 | 99.7 |
| **GBM** |  |  |  |  |  |
| VUMC | 90 | 90.2 | 52.7 | 1.9 | 99.8 |
|  | 95 | 95.1 | 40.5 | 2.9 | 99.8 |
|  | 99 | 99 | 13.1 | 4.7 | 99.7 |
| MGB | 90 | 90.2 | 63.1 | 2.3 | 99.9 |
|  | 95 | 95.1 | 45.3 | 3.3 | 99.8 |
|  | 99 | 99.1 | 15.1 | 5.5 | 99.7 |
| GHS | 90 | 90.2 | 64.2 | 2.1 | 99.9 |
|  | 95 | 95.1 | 48.8 | 3.2 | 99.8 |
|  | 99 | 99.1 | 20.4 | 6.8 | 99.7 |
| **Ridge** |  |  |  |  |  |
| VUMC | 90 | 90.1 | 50 | 1.8 | 99.8 |
|  | 95 | 95.1 | 34.8 | 2.5 | 99.8 |
|  | 99 | 99 | 9.2 | 3.3 | 99.7 |
| MGB | 90 | 90.2 | 63.6 | 2.3 | 99.9 |
|  | 95 | 95.1 | 43.9 | 3.2 | 99.8 |
|  | 99 | 99 | 10.4 | 3.8 | 99.7 |
| GHS | 90 | 90.1 | 48.8 | 1.6 | 99.8 |
|  | 95 | 95.1 | 35.1 | 2.3 | 99.8 |
|  | 99 | 99 | 10.5 | 3.5 | 99.7 |
| **Ensemble** |  |  |  |  |  |
| VUMC | 90 | 90.2 | 58.2 | 2.1 | 99.8 |
|  | 95 | 95.2 | 46.9 | 3.4 | 99.8 |
|  | 99 | 99.1 | 18.9 | 6.8 | 99.7 |
| MGB | 90 | 90.2 | 56.4 | 2.1 | 99.8 |
|  | 95 | 95.1 | 39.7 | 2.9 | 99.8 |
|  | 99 | 99 | 12.4 | 4.5 | 99.7 |
| GHS | 90 | 90.1 | 52.5 | 1.8 | 99.8 |
|  | 95 | 95.1 | 36.3 | 2.5 | 99.8 |
|  | 99 | 99 | 14 | 4.9 | 99.7 |

### Predictor Importance

The most important predictors to each model are listed in the eSupplement. Of note, importance for the tree-based methods (GBM and RF) were defined by purity, the variance in the responses with the addition/subtraction of those predictors. For Ridge, importance was defined as the magnitude of regression coefficients by predictor. The top fifty features per algorithm ranked by importance are shared in an eSupplement with the top ten for each algorithm shown here, ranked (Table 4). Ridge trained at MGB was driven by medication use while GBM at GHS and RF at VUMC was driven by demographics and diagnostic codes. We caution overinterpretation of such predictor weights and underline these statistics are correlative, not causal.

**Table 4:**
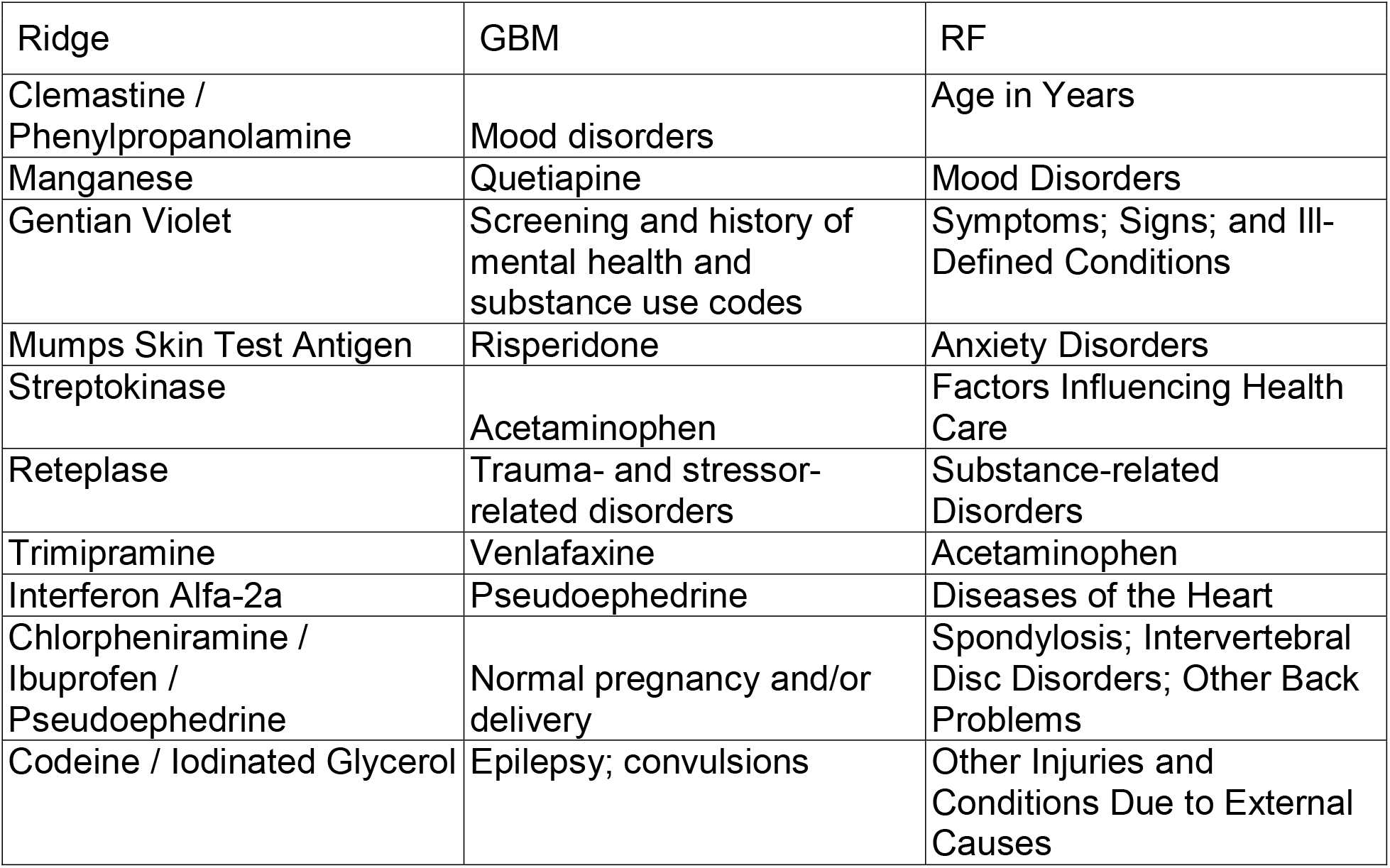
Ten most important predictors by algorithm. Importance refers to impact on model performance and confers insight into correlation, not causation.

## Discussion

Early identification of individuals at risk for BD offers opportunities for targeted assessment and prevention. Although a number of risk factors for BD have been established including family history^25^ and stressful life events^26^, quantitative, scalable prediction of risk is challenging. Prior studies have largely focused on individuals with a history of depression and/or have included relatively small samples.^27,28^ Here, we validated multiple algorithmic approaches across multiple well-powered longitudinal EHR sites in the absence of a common data model to generate a novel suite of prediction algorithms for BD. These models performed well across diverse geography and broad, heterogeneous patient populations. However, difficulty in portability and transferring algorithms across sites remains a primary barrier to replicative and implementation studies.

Our results demonstrate the feasibility and comparative performance of prediction algorithms using federated analyses of EHR data across the PsycheMERGE network. We compared three different machine learning approaches, each reliant on different assumptions and means of handling noisy, high dimensional data. Finally, we tested ensembles of these methods via stacking.

We highlight several noteworthy findings. First, we found that, regardless of method, performance was optimal at the site at which the model was developed, supporting the inference that portability of models may be limited by site-specific features - e.g., a local care practice common in one setting or region and uncommon in another. It also suggests potential for overly optimistic performance estimates with internal validation - underlining again that no substitute exists for external validation in model evaluation. We also note little overlap to the most important predictive features at each site, which likely relates to both site-specific differences and algorithmic differences, e.g., parametric (Ridge) and nonparametric (tree-based) approaches. The most generalizable algorithm, the stacked ensemble, matched internally valid algorithms in discrimination and was the only well-calibrated model, in part owing to its recalibration via regression at each site during the stacking process. Vigilance for drift and miscalibration over time would be necessary in planning implementation downstream.

Race was included as a predictor in these models, a feature being reconsidered for a number of clinical predictive applications. We opted not to blind our algorithms in this case as this process has been shown not to prevent algorithmic bias and might, in fact, introduce it. We emphasize race is a social construct that does not itself cause mental illness but can be a marker of inequitable healthcare access, experiences of adversity, and systemic inequity of opportunity. As such, it might be predictive of a coded diagnosis despite not being in the causal path for the outcome. Prior to implementation of models like these, close attention to algorithmic bias and potential for disparities should be considered using variables like coded race for prediction.^29,30^ As an additional check that race as a predictor did not bias our model unfairly, we retrained the RF at VUMC and compared validation set performance by coded race across 1,000 bootstrap replicates. Performance distributions did not differ across or within race: coded White race AUC 0.8 [0.79, 0.82] and 0.79 [0.77, 0.8] for model with and without race, respectively; coded Black race AUC 0.79 [0.76, 0.83] and 0.78 [0.75, 0.82] for model with and without race, respectively.

The clinical utility of predictive models for rare events like BD (<1% at each site) merits consideration and attention to the importance of PPV. Here, the stacked ensembles achieved the best threshold-specific PPVs across all three sites (a ∼forty-fold increase compared to case prevalence). A resource-limited clinical environment prioritizing identifying those most likely to have undiagnosed BD or predicting onset of BD might benefit from models that provide such PPVs in the setting of such rare outcomes. Of note, our models achieved a NNS as low as 20 or fewer at each site, meaning that fewer than 20 patients would be identified as high risk for every true case detected.

These models rely on EHR data common to any modern hospital agnostic to a common data model: demographics, diagnostic codes in a universally accepted schema (ICD), medications mapped to a public ontology (RXNORM). For those who wish to leverage these trained and tested algorithms, a library of the individual models and the stacked ensemble will be made available on the PsycheMERGE Consortium website (psychemerge.com).^31^

### Strengths

This study leveraged three large health systems with a validated definition of bipolar disorder as the prediction target. We applied three accepted algorithms (RF, Ridge Regression, GBM) to large real-world cohorts and assessed generalizability and model fit across partner sites. We ensembled these algorithms on over 3M patient lives across three major biobanks - the largest modeling study of this kind in BD, to our knowledge. We relied on readily available structured EHR data for feature engineering. Finally, we disseminate these tools via the PsycheMERGE Consortium to facilitate replication studies and local deployment.

### Limitations

Our results should also be interpreted in light of several limitations. First, while we explored performance of multiple different modeling approaches, there are others (including deep learning approaches) that were not tested. Second, our study relied on structured longitudinal EHR data, a decision we made to facilitate ready implementation across sites. However, natural language processing of narrative text might offer performance advantages longer term. Third, covariate shift in real-world data like these mean the joint distribution of model inputs and outputs may differ between training and testing across sites.^32^ Methods of covariate shift detection and adaption might be investigated using importance re-weighting or feature dropping methods in future studies to improve model performance. Finally, class imbalance remains a notable challenge in this study and studies like it, and creates the potential for overfitting and spuriously high model performance metrics (e.g., high AUROCs simply because of identification of the majority class, here non-BD).

## Conclusions

Generalizable predictive models of bipolar disorder trained and validated across health systems are feasible targets of clinical and precision medicine focused initiatives, even in the absence of common data models across sites. Implications of these models include BD risk research acceleration, catalyzing pharmacoepidemiologic studies, and potential for similar models to serve as probabilistic phenotypes in precision medicine research. Future work should assess their clinical utility and potential to phenotype quantitatively this serious mental illness.

## Data Availability

Study data including de-identified electronic health records linked to biobanks. However, complete anonymization to prevent inadvertent or intentional reidentification is not possible with granular healthcare data as those used here. Study-related analytic code and trained algorithms will be made available with publication as per the manuscript text.

## Acknowledgments

All investigators were supported in part by NIMH R01MH118233 (PIs Smoller/Davis). Dr. Smoller is also supported in part by a gift from the Ryan Licht Sang Bipolar Foundation. Dr. Davis is also supported in part by R56MH120736. Dr.

Walsh is also supported in part by NIMH R01MH121455 and R01MH116269. Dr. Choi is supported in part by a NARSAD Young Investigator Grant from the Brain & Behavior Research Foundation.

Funders played no role in design and conduct of the study; collection, management, analysis, and interpretation of the data; preparation, review, or approval of the manuscript; and decision to submit the manuscript for publication.

## Author Contributions

Design and conduct of the study (all authors)

Data collection, management, analysis (C.W., Y.H., M.R., D.R., D.W., Y.S., K.C., A.Z.) Interpretation of the data (all authors)

Preparation, review, or approval of the manuscript (all authors)

## Competing Interests

Authors (Ripperger, Hu, Sheu, Wilimitis, Rocha, Choi, Castro, Kirchner, Chabris, Davis) declare no Competing Financial or Non-Financial Interests

Dr. Smoller declares no Competing Non-Financial Interests but the following Competing Financial Interests: PI of a collaborative study of the genetics of depression and bipolar disorder sponsored by 23andMe for which 23andMe provides analysis time as in-kind support but no payments. He is also a member of the Scientific Advisory Board of Sensorium Therapeutics (with equity).

Dr. Zheutlin declares no Competing Non-Financial Interests but the following Competing Financial Interests: currently employment by Janssen Pharmaceuticals. Other than manuscript revision, she contributed to this research only during her time as a postdoctoral fellow at MGB.

Dr. Walsh declares no Competing Non-Financial Interests but the following Competing Financial Interests: equity interest in Sage AI, LLC (unrelated to healthcare).

## Supplemental Material

**Table S1:** Clinical variables considered in developing predictive models for Bipolar Disorder

| *Feature Category* | *# of Variables (VUMC)* | *# of Variables (MGB)* | *# of Variables (GHS)* | *Details* |
| --- | --- | --- | --- | --- |
| *Demographics* | *3* | *3* | *3* | *Age, Sex, Race* |
| *Comorbidities* | *135* | *130* | *130* | *All comorbidities are mapped to CCS code* |
| *Medications* | *2367* | *1712* | *1134* | *All medications are mapped to RxNorm ingredient* |

## Notes

### Competing Interest Statement

The authors have declared no competing interest.

